# A mixed cross-sectional and case-control study approach to investigate the risk-factors of Noma/Cancrum oris in Ethiopia

**DOI:** 10.1101/2022.03.31.22273219

**Authors:** Heron Gezahegn Gebretsadik, Laurent Cleenewerck de Kiev

**Affiliations:** School of Global Health & Bioethics, Euclid University, Banjul, Gambia; EUCLID (Euclid University), Bangui, Central African Republic

## Abstract

**Introduction:** Noma is a polymicrobial gangrenous facial disease affecting people living in the most impoverished areas of low- and middle-income countries. If left untreated, the disease is fatal or else severely disfigure people with the condition. The compromised immune system, poor oral hygiene, measle infection, diarrheal disease, inaccessibility to health education and proper medical care, and lack of a balanced diet and good sanitary facilities are found to be some of the predisposing factors for the development and progression of the disease. Furthermore, debilitating diseases like malaria and measles were considered as significant precursors to Noma.

**Materials and Method:** A mix of cross-sectional and case-control study approaches was conducted to assess the risk factors of Noma in Ethiopia. The raw data of the cases were obtained from Yekatik 12 Hospital, Facing Africa, and the Harar project Ethiopia. Three controls were selected per single case. The Odd ratio (ORs) and Chi-square test were calculated to rule out the statistical significance of the association observed between the factors and the disease.

**Results:** A total of 64 cases were selected for the case-control study. Considering the 1:3 case to control ratio, 192 matching controls were identified. Malaria, helminths, measle, diarrheal diseases, and living with domestic animals were found to be risk factors for Noma with a respective p-value < 0.01. Contrarily, the analysis has identified vaccination (p < 0.01) as a protective factor.

**Discussion:** Noma/face of poverty is mostly preventable by providing proper nutrition, sanitary and water facilities, awareness about the disease, oral health education, and vaccinations. Poverty-related diseases such as malaria, helminths infection, measle, diarrheal diseases, and unfavorable living conditions were identified to be the risk factor for Noma. As such the disease is truly preventable. Prevention of the disease can be achieved through promoting overall awareness of the disease, poverty reduction, improved nutrition, and promotion of exclusive breastfeeding in the first 3-6 months of life. Furthermore, optimum prenatal care, timely immunizations against common childhood diseases, initiating vaccination, and improving the social living conditions are the other preventive mechanisms. Moreover, long-lasting economic development should be considered to effectively and sustainably prevent the disease.

## Introduction

Noma is a rapidly progressive, polymicrobial, opportunistic, gangrenous infection of the mouth, most likely caused by certain kinds of Burulibacterial flora that switch to pathogenic when the host is immunocompromised (1). Alternatively known as Stomatitis gangrenosa or Cancrum oris, the etiology of Noma is infectious, yet unclear as regards the exact causative microorganism (2). Due to its primary occurrence in young children living in remote areas of the least developed countries with inadequate health systems and its rapid progression rate and high case fatality, today, Noma is a poorly studied disease, and its cause remains idiopathic (3). Because of the potential involvement of bacteria, the transmissibility of Noma has to be considered, but epidemiological patterns and animal experiments currently do not support this possibility (4).

Several potential pathogens were found in abundance in the sites of Noma, which include Prevotella melaninogenica, Corynebacterium pyogenes, Fusobacterium nucleatum, Bacteroides fragilis, Bacillus cereus, Prevotella intermedia, and Fusobacterium necrophorum. Microbial analysis in the early 20th century revealed the presence of spirilliform and fusiform microorganisms in biopsy samples taken from the transitional zone between gangrenous and healthy tissues (5). Later studies reported that Fusobacterium necrophorum, a predominant animal pathogen is the most common microorganism isolated from the disease sites in Nigerian children. It was suggested that Fusobacterium necrophorum could be a trigger organism for noma. This microorganism produces various toxins and has been associated with necrotizing infections in animals, and it may contaminate livestock and potentially infect children (6).

On the other hand, a recent study has contradicted the involvement of Fusobacterium necrophorum as an etiologic agent (7). Known periodontal pathogens like aggregatibacter actinomycetemcomitans, capnocytophaga, porphyromonia, and fusobacteria were more prevalent in healthy samples compared to those with Noma (8). The study also identified prevotella intermedia and peptostreptococus to be more clearly associated with Noma (9).

Noma is commonly seen in a population with extreme poverty, severe malnutrition, unsafe drinking water, poor sanitation, poor oral health practices, limited access to high-quality health care, and intrauterine growth retardation (10). Recently, an increased incidence of Noma has been reported in patients with Human Immune deficiency Virus (HIV) infection, cyclic neutropenia, leukemia, Down’s syndrome, Burkett’s disease, and herpetic stomatitis (11).

Malnutrition is considered to be an important risk factor for Noma (12). In Africa, most of the cases were reported during the dry season when food is scarce, and when the incidence of measles is highest. Measles could also be an important risk factor because of its associated immunosuppression (13).

A retrospective study revealed severe malnutrition, recent respiratory or diarrhoeal syndrome, the number of previous pregnancies in the mother, an altered oral microbiota, and the absence of chickens at home as predictors of Noma (14). Furthermore, lack of maternal care, measles, and malnutrition were reported as risk factors for developing Noma in Nigeria (15) (14).

In general, immunodeficiency disorders, including AIDS, down syndrome, malnutrition, dehydration, poor oral hygiene, recent orofacial and systemic illness, unsafe drinking water, poverty, malignancy, and living near domestic animals are reported to be other predisposing factors (16) (17) (18). The unusual case of Noma in a 32-year-old malnourished HIV-seropositive female, within three days, the initial intraoral necrotizing process spread rapidly and caused full circular thickness perforating destruction of the lower lip has shown immunosuppression associated with a high HIV load as an important risk factor in South Africa (19). On many occasions, recent debilitating conditions like measles (commonly), herpès simplex, varicella, scarlet fever, malaria, gastroenteritis, tuberculosis, and bronchopneumonia appear to set the stage for the development of Noma (1). Also, intrauterine growth retardation, down syndrome, exposure to cytomegalovirus, and premature birth have been considered as predisposing factors for the development of Noma. Cases associated with malignancies (e.g., leukemia) are not rare (18).

Mortality used to be a common complication of Noma (20). Only approximately 15% of children survive acute Noma. However, with the use of modern antibiotics and better nutrition, the mortality rate has reduced from 90% to 8-10% (21). Most Noma patients have difficulty in mastication because of the loss of soft and hard tissues. Severe cosmetic disfigurement can also take place from the resulting scarring and loss of tissue (22). At a later age, these contractures often lead to growth disturbances and result in further facial disfigurement and functional impairment. The psychological impact of surviving Noma can be easily understood; patients reported high psychiatric morbidity after Noma (23).

Noma is a truly enigmatic disease and very little epidemiological global data are available (24). The risk factors of the disease are not also well-understood (25). This explains the extent to which the disease is neglected (26). Yet, the disease affects poor children among the disadvantaged societies throughout the world including Ethiopia (27). This study was initiated to assess the risk factors of Noma in Ethiopia.

## Materials and Methods

### Study design

A mix of cross-sectional and case-control study approaches was used to identify the possible risk/predisposing factors that could potentially be associated with the development of Noma in this study. The study matched three controls for each corresponding case in terms of geographic location, gender, and age.

### Study location and population

The cross-sectional and case-control studies were conducted in different parts of the country, including Addis Ababa depending on the findings of the demographic-geographic data obtained from patients’ medical records review. All the patients’ medical records were obtained from the main offices of the three major Noma treatment centers located in Addis Ababa. Cases were patients diagnosed with Noma and treated in Yekatit 12 Hospital, Facing Africa Ethiopia or Harar Project Ethiopia between March 2004 and December 2020. The cases were identified via the cross-sectional study. Controls were individuals that matched to cases by the village of residence, current age (+/- 2 years), and sex.

### Case and control definitions

#### Definition of case

In this study, cases are individuals who have been diagnosed with Noma in Ethiopia between January 2004 and February 2020. To be ascertained as a case of Noma, an individual should have facial edema, intraoral necrotizing stomatitis, metallic taste, typical halitosis (often considered as pathognomonic), bluish discoloration of the skin, gangrenous demarcation, tissue necrosis, and sloughing. Moreover, three or more of the following should be considered: a history of malnourishment, a history of poor oral hygiene, a history of poverty, a history of poor sanitation and living close to domestic animals, and a history of debilitating and immunocompromising diseases such as measle, tuberculosis, AIDS, leukemia.

#### Definition of control

Controls are individuals who were never diagnosed with Noma. Furthermore, the controls need to be matched to cases by the village of residence, current age (+/- 2 years), and sex.

### Data collection and analysis

The investigator needed to review the medical registers of Noma cases for the 2004-2020 period including patients’ photos with relevant clinical and demographic information to answer the research questions. A modified case report form (CRF) consisting of demographic and clinical sections was used to collect the relevant demographic and clinical information, respectively (Appendix S1). The demographic section of the CRFs contains the name, gender, age, physical address (geographic location), telephone address, and year of admission of the patients. On the other hand, the clinical section of the CRFs primarily subdues the localization of Noma-induced anatomical lesions, medical history, dietary information, vaccination details, living stands, and functional impairments data.

Similarly, a questionnaire with a set of questions was used to interview the cases (caregivers or guardians) and controls (caregivers or guardians) to complete the case-control study (Appendix S2). Therefore, the information regarding the possible risk factors for Noma was obtained using a structured written questionnaire and reviewing patients’ medical charts. The questionnaire also consisted of demographic and clinical (risk factor) data. Both the CRFs and questionnaire were made to be valid. The researcher verified the validity of the CRFs and the questionnaire. Furthermore, pre-tests were conducted to assess the validity and reliability of the data collection instruments that were used in this case-control study.

On the other hand, the contact information of the cases, including patient name, telephone number, and home address, were obtained from the CRFs and used to select neighboring controls. The controls were explored for the necessary demographic and clinical information to carry out the case-control study. The data analysis of the case-control study follows the entry of relevant clinical and demographic data of the cases and controls into an excel sheet. Descriptive analysis was used to determine the characteristics of cases and controls. The odds ratio (OR) was calculated to rule out the existing association between the possible risk/predisposing factors and the disease. A Chi-square test was carried out to test the statistical significance of the associations observed between the predisposing factors and the condition.

### Ethical consideration

The Addis Ababa Health Bureau has given the necessary in-country ethical clearance. Oral consent was received from all the study participants before starting the interview to complete the questionnaire. The interviewees were fully informed about the study and its purpose, expected risks or benefits from participation, and confidentiality of information. All the information was treated secretly. Moreover, the researcher used patients’ initials in the excel sheet.

## Results

The case-control study was employed to identify the possible risk factors to develop Noma among the cases. The cases were selected from the medical records obtained from the three Noma treatment centers (Yekatit 12 Hospital, Facing Africa, and Harar Project). Completeness of medical records, availability of pertinent contact information (at least phone number), and age of the patient during admission were the criteria to select the 64 cases.

Out of the 163 Noma patients who had sought care in the programs between March 2004 and December 2020, 64 were eligible for inclusion in the case-control study. Of these, one could not be located for logistical reasons, and the researcher managed to interview the study participant via phone. Thus, the final analysis included 64 Noma cases and 192 controls. All cases and controls were under 41 years old. The age restriction was considered to avoid recall bias. The assumed potential risk factors were living with domestic animals, having diarrheal disease, having a helminthic infection, exposure to malaria infection, being inflicted with measle infection, and drinking river (Tables 1-6).

**Table 1.**
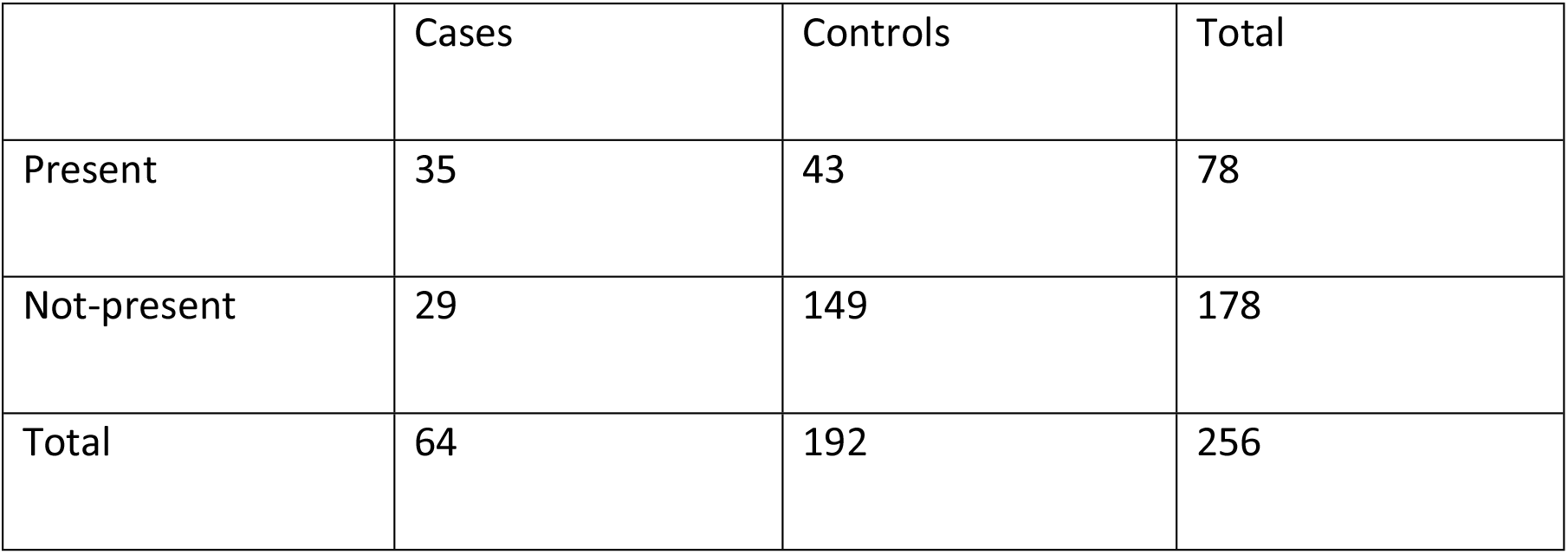
Domestic animals at home (2 × 2 table)

|  | Cases | Controls | Total |
| --- | --- | --- | --- |
| Present | 35 | 43 | 78 |
| Not-present | 29 | 149 | 178 |
| Total | 64 | 192 | 256 |

**Table 2.**
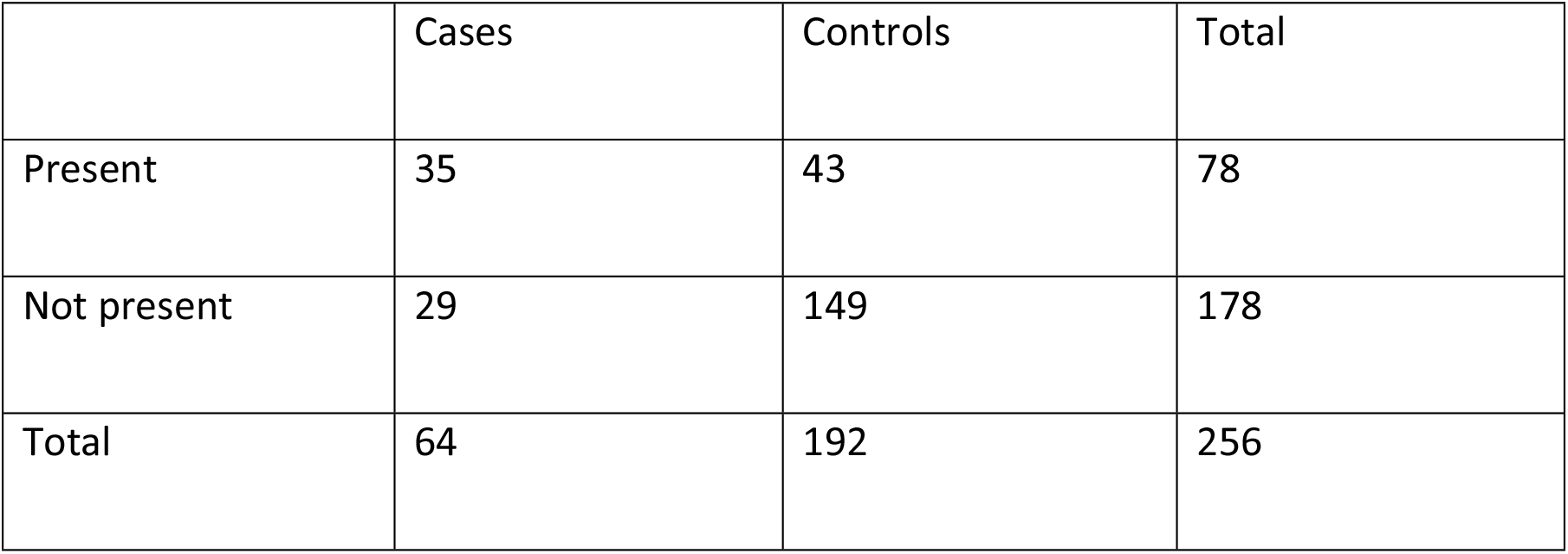
Diarrheal disease (2 × 2 table)

**Table 3.** Helminthic infection (2 × 2 table)

|  | Cases | Controls | Total |
| --- | --- | --- | --- |
| Present | 31 | 12 | 43 |
| Not present | 33 | 180 | 213 |
| Total | 64 | 192 | 256 |

**Table 4.** Malaria infection (2 × 2 table)

|  | Cases | Controls | Total |
| --- | --- | --- | --- |
| Present | 21 | 14 | 35 |
| Not present | 43 | 178 | 221 |
| Total | 64 | 192 | 256 |

**Table 5.** Measle infection (2 × 2 table)

|  | Cases | Controls | Total |
| --- | --- | --- | --- |
| Present | 13 | 14 | 27 |
| Not present | 51 | 178 | 229 |
| Total | 64 | 192 | 256 |

**Table 6.** Drinking river water (2 × 2 table)

|  | Cases | Controls | Total |
| --- | --- | --- | --- |
| Drinks | 15 | 41 | 56 |
| Not drink | 49 | 151 | 200 |
| Total | 64 | 192 | 256 |

On the other hand, vaccination was regarded as a potential protective factor based on the literature review (Table 7).

**Table 7.** Vaccination (2 × 2 table)

|  | Cases | Controls | Total |
| --- | --- | --- | --- |
| Vaccinated | 14 | 104 | 118 |
| Non-vaccinated | 50 | 88 | 138 |
| Total | 64 | 192 | 256 |

**Table 8.** Summary of risk and protective factors.

| Risk/protective factor | Odd ratio (ODs) | Chi-square ($\chi^2$) | p-value |
| --- | --- | --- | --- |
| Malaria | 6.2 | 24.37 | 0.0000008 |
| Helminthic infection | 14.1 | 58.11 | 0.0000000001 |
| Measle infection | 3.2 | 7.30 | 0.007 |
| Diarrheal diseases | 4.2 | 22.10 | 0.000003 |
| Domestic animals | 4.1 | 22.13 | 0.000002 |
| River water | 1.13 | 0.03 | 0.9 |
| vaccination | 0.24 | 21.46 | 0.000004 |

The clinical information obtained from the patients’ medical records were the main guiding principles to consider these as a potential risk and protective factors for Noma. Accordingly, each potential risk factor and the vaccination as protective factors were assessed for having a statistically significant association with the occurrence of Noma.

In conclusion, malaria, helminthic and measle infections, living with domestic animals, and diarrheal diseases are identified to be risk/predisposing factors for the development of Noma. On the contrary, drinking river water has shown no significant association with the development of Noma among the cases. On the other hand, vaccination is found to be a protective factor not to develop Noma among the controls.

## Discussion

The vast majority of the Noma cases are living in the most deprived and remote regions of sub-Saharan Africa (28). Historically, Noma was most prevalent in the Noma belt region that extends from Senegal to Ethiopia (5). Studies have shown that in countries where the condition is widespread, risk factors such as malnutrition, debilitating diseases like malaria and measles, close residential proximity to livestock are reported (29) (30). Moreover, respiratory or diarrheal syndrome and an altered oral microbiota have been reported as risk factors of Noma (14). The HIV/AIDS pandemic increases the number of cases outside the Noma belt region and is considered to be a viable risk factor of the disease (31). Other risk factors that have been reported in several articles include the absence of breastfeeding, unsafe drinking water, limited access to high-quality health, and food security (1) (32) (33).

On the other hand, childhood vaccine coverage has been reported as a protective factor not to develop Noma (34). Diarrheal diseases, drinking river water, living with domestic animals, vaccination coverage, and measle, malaria, and helminthic infections were tested for possible significant association with the development of Noma in this study. The findings of this study supported the above argument. In other words, measles, malaria, helminthic infections, diarrheal diseases, presence of domestic animals at home have been identified as potential risk factors to develop Noma among the cases. On the other hand, vaccination coverage has shown a statistically significant protective effect not to develop Noma among the controls.

Malnutrition is considered to be an important risk factor for Noma (35) (36). In Africa, most of the cases were reported during the dry season when food is scarce, and when the incidence of measles is highest (37). The debilitating diseases such as malaria and measles have shown a statistically significant association of being precursors to Noma in this study. Hence the periodicity of onset of the disease in this study could be explained at the dry seasons as food is scarce, and the incidence of measle and malaria often increase (38). Measles and malaria could also be important risk factors because of the associated immunosuppression (39) (40). Furthermore, malnutrition could be regarded as a confounding factor in these associations. However, a further in-depth investigation needs to be carried out.

The proportion of vaccination coverage in many developing countries is below the standards recommended by the WHO (41). There is evidence that the occurrence of vaccine-preventable diseases and malnutrition precedes the onset of Noma. The low coverage of vaccination not only increases the risk of morbidity and mortality from vaccine-preventable diseases but is a contributing factor in immunosuppression, which is thought to play a vital role in the sequence of events for Noma development (42). Measle, the only vaccine-preventable disease that has been identified as a risk factor of Noma in this study, could cause immunosuppression among the cases described (43). The diarrheal and helminthic infections that have been identified in this study could also cause immunosuppression. This immunosuppression could be generally confounded by low coverage of vaccination.

On the contrary, the protective effect of vaccine coverage that has been identified in this study could be regarded as a positive factor to reduce the burden of the disease. Living with domestic animals is found to be the other risk factor of Noma among the cases studied in this study. This factor could be explained in terms of the lack of proper sanitation. On the other hand, drinking river water has shown neither protective nor causative effect on the development of Noma in this study.

The tropical climate, lack of education, rural condition, poor sanitation, and poverty are the leading causes of the high occurrence of Noma (11). A tropical climate characterizes the Afar, Somali, Gambella, Tigray, part of Amhara and Oromia regions. These regions constituted nearly 73% of the total cases admitted to the three Noma treatment facilities in Ethiopia. Climate could be also considered as a potential risk factor for the development of Noma in Ethiopia. Nevertheless, further investigation needs to be carried out.

Furthermore, several studies have shown that in countries where the condition is widespread, risk factors such as malnutrition, debilitating diseases like malaria and measles, close residential proximity to livestock are reported (44) (45). Moreover, respiratory or diarrheal syndrome and an altered oral microbiota have been reported as potential risk factors of Noma. The HIV/AIDS pandemic increases the number of cases outside the Noma belt region and is considered to be a viable risk factor of the disease (46) (47). On the other hand, childhood vaccine coverage has been reported as a protective factor for developing the condition. Similarly, in this study measles, malaria, and helminthic infections, diarrheal diseases, and the presence of domestic animals at home have been identified as potential risk factors to develop Noma among the cases. On the other hand, vaccination coverage has shown a statistically significant protective effect not to develop Noma among the controls.

In general, the risk factors of Noma reported in the contemporary literature are highly related to poverty (48). In other words, Noma can be considered as an excellent biological parameter of extreme poverty. Hence, policymakers need to be aware that activities against extreme poverty can inevitably avert the occurrence as well as an ill sequela of the disease in developing countries (27).

Noma is a disfiguring necrotizing disease of the Oro-facial tissue. It is an “infectious” disease of unknown etiology (49). The burden of the disease can be explained through its high mortality rate and psychosocial morbidity (50). Following the devastating facial disfigurements, survivors are often discriminated against from social life (51). Noma survivals are often left with devastating orofacial defects, which resembles a dramatic amalgamation of oncologic, congenital, and traumatic deformities (52). For instance, a retrospective study that included 22 Noma cases who undergo surgical intervention in Nigeria has revealed 83.3% had lip involvement (53).

## Conclusions

In conclusion, this study suggests that disadvantaged children under the age of 10 are predominantly affected by the disease. Most of the victims have shown remarkable Noma-induced facial disfigurements, which exposed them to social discrimination and tangible psycho-social crisis. Living with domestic animals, inaccessibility of adequate vaccination coverage, high morbidity of infectious diseases such as measles, malaria, helminths, and diarrheal diseases have been identified as potential risk/predisposing factors for Noma in Ethiopia. Debilitating conditions, along with malnutrition, can explain the overall risk factors identified in this study. Though adequate data on risk factors are still lacking, the information obtained from this study underscores that Noma must not be neglected.

Noma is a debilitating disease primarily affecting the poor and has been called the “face of poverty.” It occurs among the underprivileged society where patients do not have access to proper medical care. Poverty, along with low access to health care, can explain the overall risk factors identified in this study. In other words, Noma can be considered as an excellent indicator of extreme poverty. Improving the economic status of the disadvantaged communities can be considered as the crucial strategy to eradicate Noma globally. History has revealed that Noma can be completely eradicated by enhancing economic progress and reducing poverty. Therefore, strong socio-economic and health policies need to be developed to enable the poorest to afford to acceptable life and access medical care (preventive and surgical rehabilitation), respectively.

## Data Availability

All data produced in the present work are contained in the manuscript

## Acknowledgment

I would like to acknowledge with gratitude the School of Global Health and Bioethics at EUCLID (Pôle Universitaire EUCLIDE) for supporting the project with useful scientific advice.

## Appendices

### Appendix S1: Case Report Form

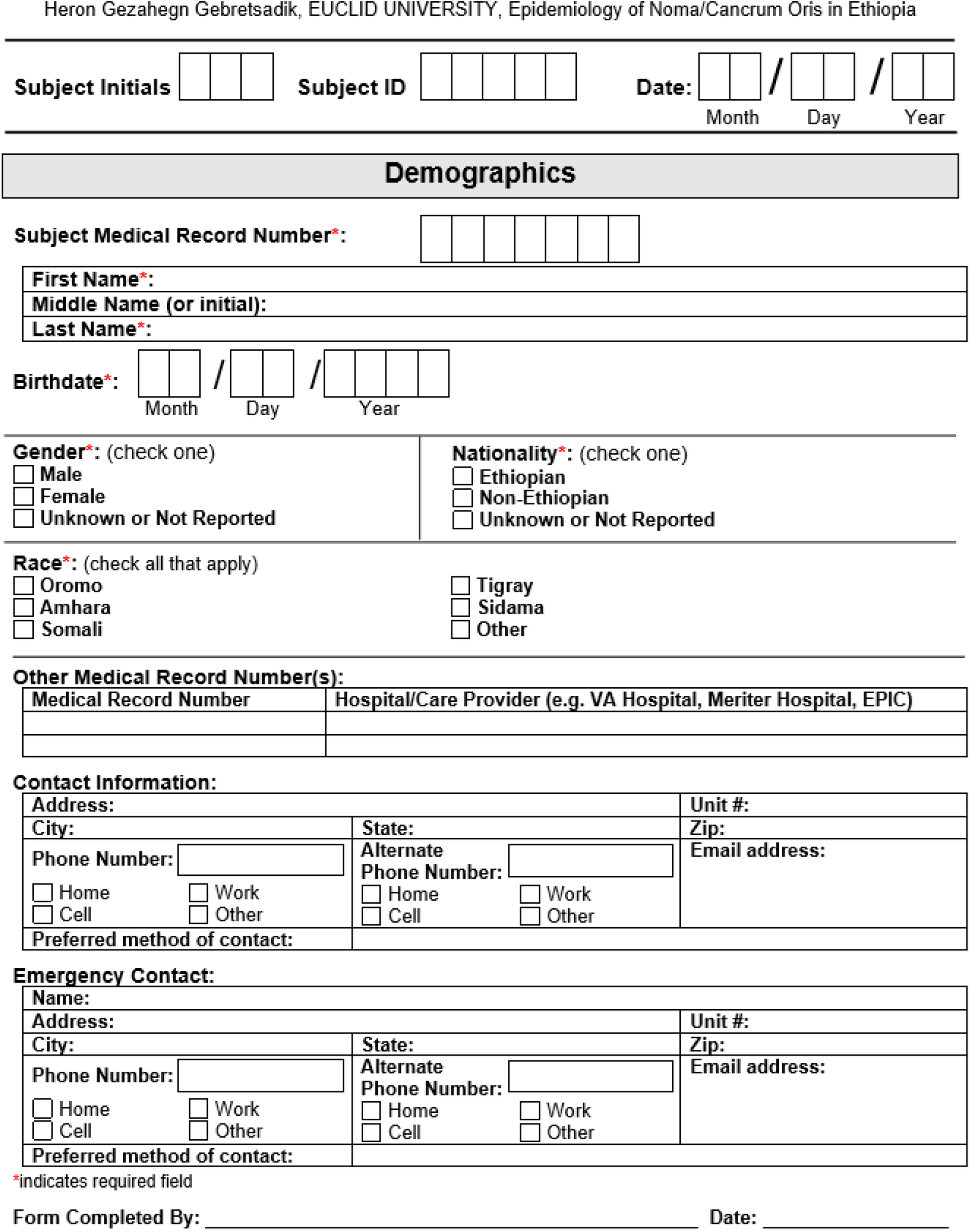

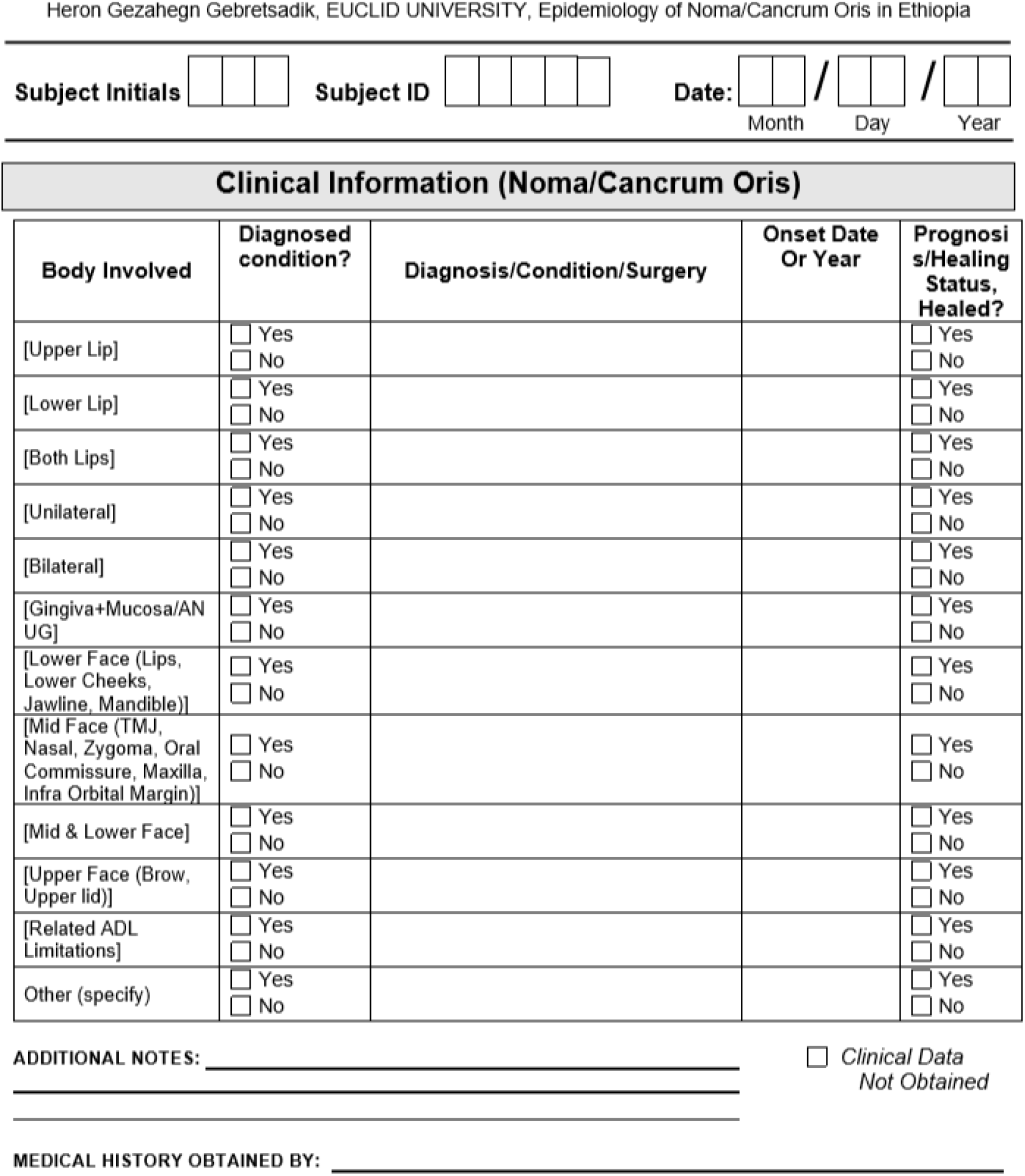

### Appendix S2: Questionnaire

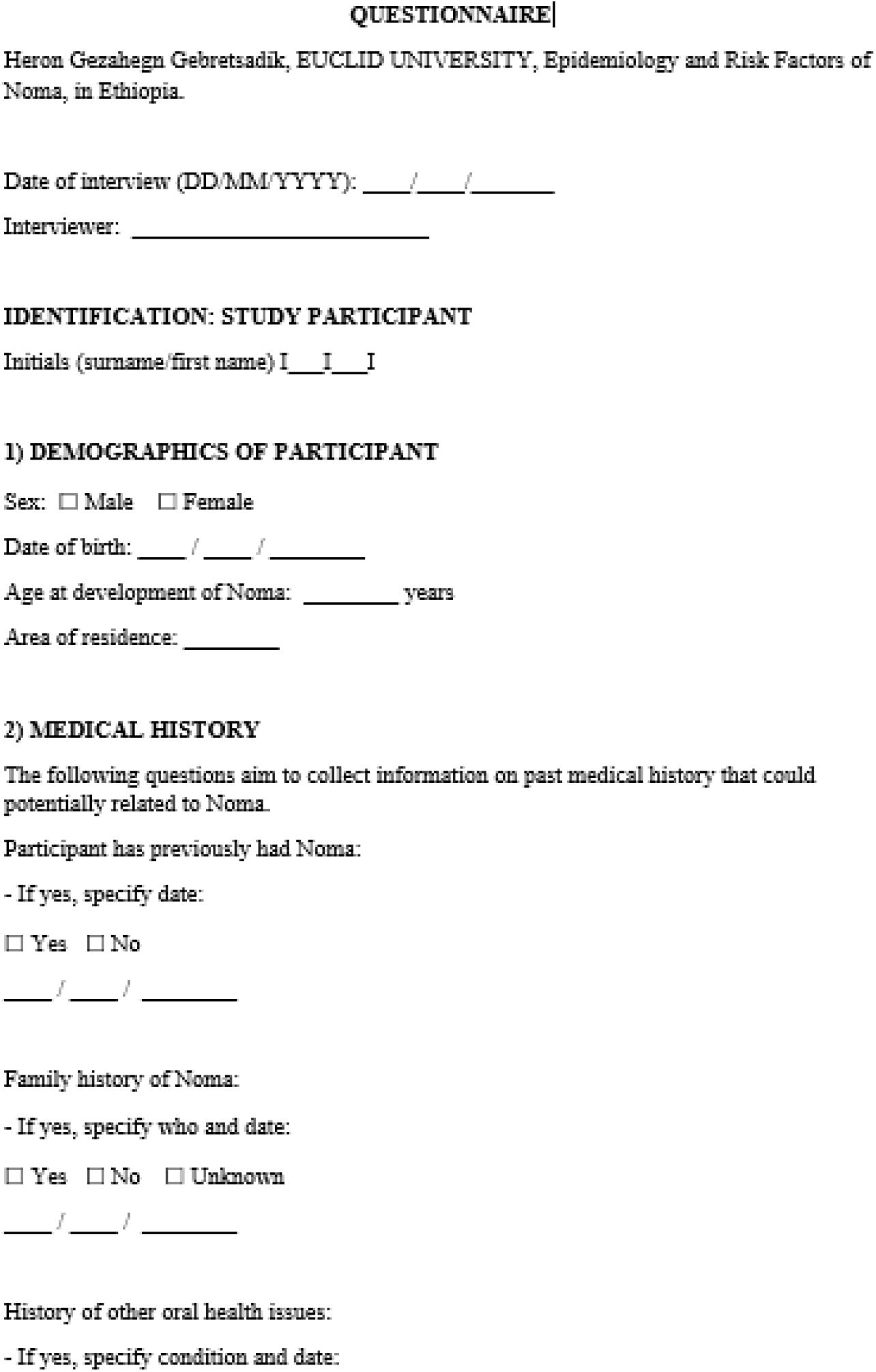

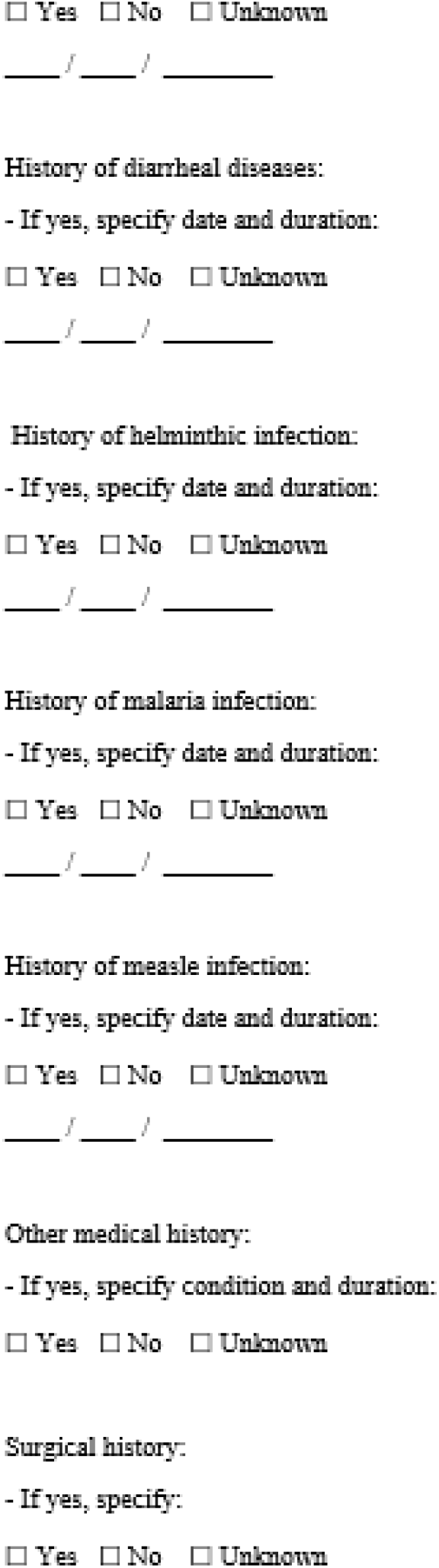

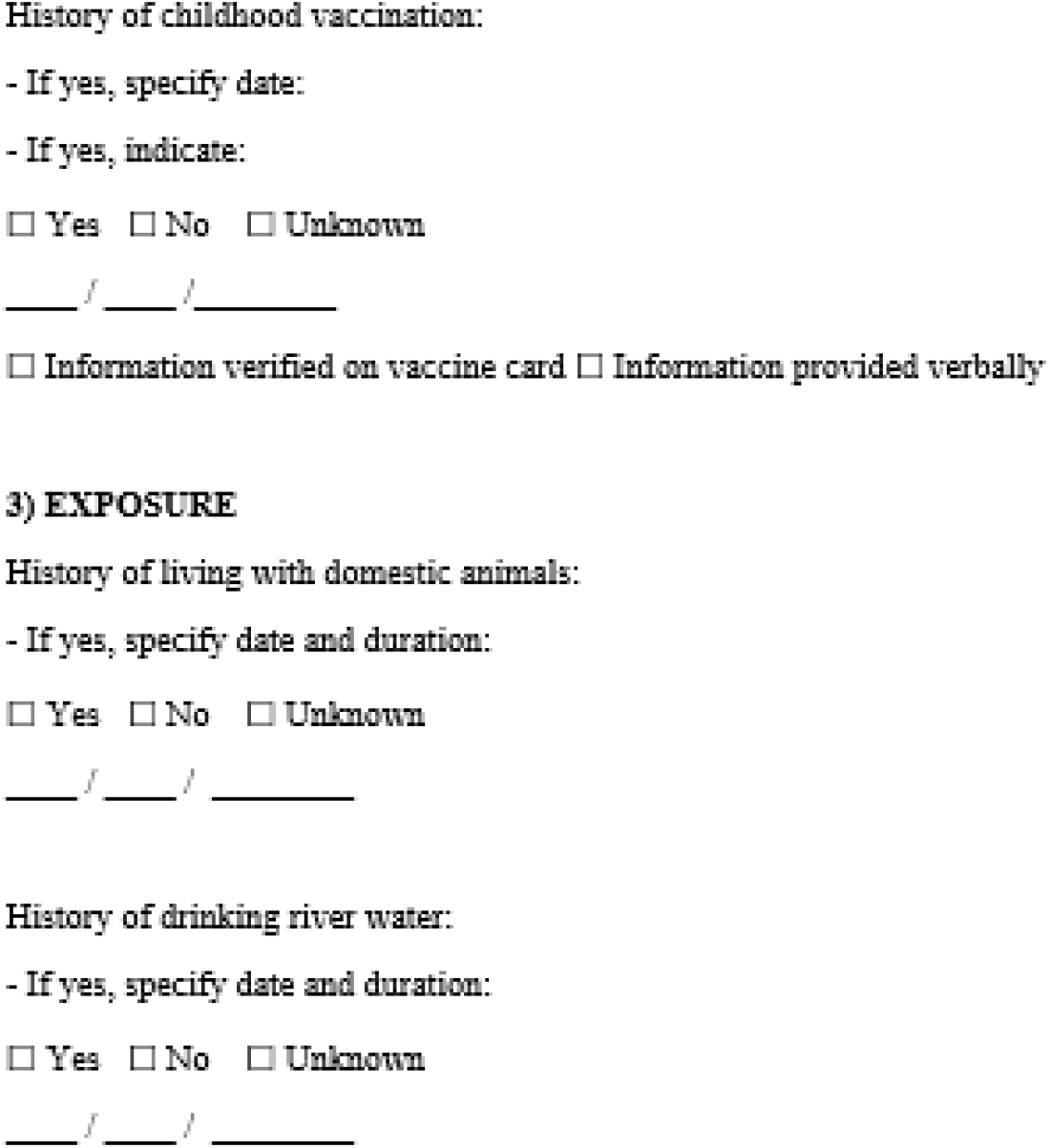

